# National Health Insurance Coverage and COVID-19 vaccine acceptance in Uganda. Implications on Uganda’s achievement of Universal Healthcare Coverage and Sustainable Development Goals

**DOI:** 10.1101/2022.08.09.22278595

**Authors:** Judith Aloyo, Freddy Wathum Drinkwater Oyat, Lawence Obalim, Eric Nzirakaindi Ikoona, David Lagoro Kitara

## Abstract

**Background:** With the advent of the novel coronavirus disease (COVID-19) and the severe second wave that caused high-profile deaths, hospitalization, and high treatment costs in Uganda, the population has raised concerns about the enactment of the national health insurance coverage bill.

As of March 31, 2021, when Uganda was beginning to experience the second wave of COVID-19, the Parliament of Uganda passed a national health insurance bill that outlined the general structure for the first national health insurance scheme. The bill had pre-set benefit packages including a wide range of essential health services such as family planning, vaccination, and counseling. The plan was proposed to be financed by a combination of employers and government contributions and aimed to cover all Ugandans when fully implemented. The policy and implementation details would evolve when the President enacts it into law. However, the President has not assented to the bill.

This study aimed to determine the prevalence of health insurance coverage and factors associated with COVID-19 vaccine acceptance among participants in northern Uganda and use findings to show its implications on Uganda’s achievement of Universal Health Coverage and Sustainable Development Goals.

**Methods:** We conducted a cross-sectional study among seven hundred and twenty-three adult participants from northern Uganda. Participants were selected randomly and consecutively. We used a questionnaire with an internal validity of Cronbach’s a=0.772 to collect quantitative data from participants. A local IRB approved the study, and we used SPSS version 25.0 for data analysis. A p-value less or equal to 0.05 was considered significant.

**Results:** The prevalence of health insurance coverage among the study population was low, 57/723(7.9%), with most insured 42/57(73.7%), accepting the COVID-19 vaccine with a mean age of 33.81 years SD+8.863 at 95% CI:31.46-36.16 and a median age of 35 years. Participants without insurance coverage but accepted the COVID-19 vaccine were 538/723(74.4%) with a mean age of 31.15 years SD+10.149 at 95% CI:30.38-31.92 and a median of 29 years. The insured and uninsured ages range from 18-52 years and 18-75 years, respectively. COVID-19 vaccine acceptance was higher among the insured 42/57(73.7%), and the likelihood ratio for insured participants to accept than reject the COVID-19 vaccine was 9.813; df=4; p=0.044. Widows, divorcees, and married separate, participants from remote districts (Nwoya and Lamwo), and those without formal education had no health insurance cover. However, in a multivariable logistic regression analysis, health insurance coverage was not an independent predictor of COVID-19 vaccine acceptance AoR=1.501,95%CI:0.807-2.791; p=0.199.

**Conclusion:** As the world grapples with the control of COVID-19, vaccine acceptance and health insurance coverage have become critical issues to be handled by each country. The health insurance coverage among participants from northern Uganda was low at 57/723(7.9%). Most participants with health insurance coverage accepted the COVID-19 vaccines compared to those who did not. The lack of health insurance coverage among most study participants is problematic as the world looks toward attaining UHC and SDGs. We proposed that Uganda’s national social health insurance scheme, which is not legal, is urgently reviewed and signed to allow Uganda’s population access to the needed health services.

## Introduction

According to the World Bank (WB) and the World Health Organization (WHO), at least half the world’s population can’t access essential health services [1]. Nearly one hundred million people worldwide are being pushed into extreme poverty by healthcare costs, meaning that after paying for vital health services, procedures, and medications, they have less than $1.90 a day to live on [1,2]. A lack of access to quality, affordable health care isn’t a problem in developing countries or countries with a high poverty rate, and it is an issue that people worldwide, from the United States of America to Chad to China, are dealing with [1]. But a world where there are adequate and affordable health services is not impossible to achieve [1]. Evidence from countries working to achieve universal health coverage (UHC), the dream of health care for all is already within sight [1,2]. Every United Nations (UN) member state has committed to achieving UHC by 2030 as part of the Sustainable Development Goals (SDGs), but not all have started taking the steps necessary to make healthcare services accessible and affordable within the next 12 years [1,2]. The greatest obstacle to establishing UHC is a lack of political will but as said, where there’s a will, there’s a way [1,2].

Since 2000, many countries, including Canada, Saudi Arabia, Rwanda, Cuba, Indonesia, and Kenya, have begun implementing reforms to establish UHC [1]. The reform initiatives of Rwanda and Indonesia prove that countries don’t have to be wealthy to provide affordable healthcare coverage [1]. In addition to overcoming a lack of political will, several other significant barriers to countries’ hope to achieve UHC by 2030 must be tackled [1,2]. These include a lack of trained healthcare workers, vaccines, medication, equipment, and infrastructures.

In some countries, a lack of infrastructure means that the population will travel long distances to receive the needed health services, which can be costly and, at times, dangerous to reach healthcare services and facilities [1,2]. The lack of infrastructure does not only mean individuals may be discouraged from getting preventative treatment like vaccines but also means they will wait until their lives hang in the balance to seek medical attention [1,2]. A lack of vaccines and medication implies that even when they are willing and able to seek care, treatment may not be available or affordable, or there may not be enough skilled workers to provide care [1,2]. In the past 16 years, despite its low-income economy, Rwanda has provided healthcare coverage to about 90% of its population [1]. It managed to do so by, passing policies that direct the use of tax revenue and foreign aid to cover healthcare costs and asking its citizens to pay voluntary premiums scaled by income (the New York Times reported) [1].

In Uganda, the situation is somewhat different. It is the only country in East Africa that has not enacted a national health insurance scheme but has one of the region’s highest out-of-pocket health costs [3]. An estimated 38% percent of Uganda’s health expenditures are paid by individuals through out-of-pocket expenses, followed by development partners (41%), the government (16%), and others (5%) [4]. Uganda’s current health insurance options are an employer or community-based schemes and are estimated to cover less than 2% of the population [5]. Health insurers only contribute around 1% to health spending in Uganda [6].

In addition, Uganda’s national insurance scheme, which is not yet legal, will allow the insured clients to receive information and services in both public and private sectors, increasing accountability for providers to offer competitive and high-quality services.

COVID-19 has dramatically interrupted the lives and livelihoods of many communities in Uganda, especially during the second wave, which swept across the country with thousands of high-profile deaths, hospitalization, and high treatment costs unaffordable to many communities. This problem was made worse by the high COVID-19 vaccine hesitancy and curiosity among the Ugandan communities.

This study aimed to determine the prevalence of health insurance coverage and factors associated with COVID-19 vaccine acceptance among participants in northern Uganda and to use it to explain its implications on Uganda’s achievement of Universal Health Coverage (UHC) and Sustainable Development Goals (SDGs).

## Methods

We conducted a cross-sectional survey in northern Uganda between March and April 2022 in twenty-four health facilities of the Acholi sub-region. We selected the health centers based on their participation in offering COVID-19 vaccination to the region’s population. We recruited participants (adults/>18 years) admitted or clients to outpatient clinics of health facilities in northern Uganda’s nine districts of the Acholi sub-region and had consented to the study. We excluded those who were critically ill and were unable to answer research questions. We calculated the sample size using the Raosoft sample size calculation in which the computation is on a 50% response distribution, 5% margin of error, and 95% Confidence Interval. We used this online software foundation because it is a widely utilized descriptive study formula for sample size estimation [7,8]. The research team chose this software calculator because Raosoft, Inc. form and survey software comprise a database management system of great strength and reliability that communicates with other proprietary formats. Raosoft database is a highly robust, proven system with high data integrity and security [7,8].

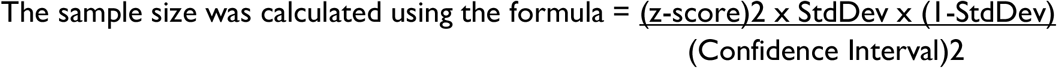

Based on a population size of 45,000 clients and OPD visitors in one month in all health facilities in the Acholi subregion, a minimum sample size grounded on the above assumptions and factoring in a 10% non-response rate is 396 participants. We used a simple random sampling technique to recruit participants. We chose this sampling technique because it is one of the most popular and simple data collection methods in research fields (in terms of probability, statistics, and mathematics). It allows for a balanced data collection, aiding studies to arrive at unbiased conclusions. The dependent variable was COVID-19 vaccine acceptance (“Have you received a jab of COVID-19 vaccine? And the answer was either “Yes” or “No”). The independent variables were the socio-demographic characteristics: age, sex, occupation, religion, level of education, tribe, marital status, district, presence of comorbidities, nationality, race, health insurance coverage, and participants’ self-confidence that the vaccines available in health facilities in northern Uganda were safe.

Our research team used face-to-face questionnaire interviews, strictly following Uganda’s standard COVID-19 infection, prevention, and control (IPC) guidelines. The questionnaire was constructed in English, consisting of questions on socio-demographic characteristics and views on the safety of vaccines in health facilities in the sub-region (Additional file 1). The questionnaire was developed and grounded on literature reviews and discussions with the research team [9,10], pretested in a regional hospital and had an internal validity of Cronbach’s α= 0.772. We assured participants’ confidentiality and privacy of their responses to reduce the potential bias introduced by self-reported data. In addition, the questionnaire was short and precise and thus minimized lethargy in participants’ responses. We conducted data analysis using SPSS statistical software version 25.0, where continuous variables were in means, standard deviations, medians, and interquartile ranges depending on the distribution of the data. Categorical data were in frequencies and percentages. The Chi-square and crosstabs tests were performed on categorical data when comparing two or more groups. In addition, we conducted a multivariable logistic regression analysis to identify independent factors associated with the COVID-19 vaccine acceptance among the insured participants and relationships between independent and dependent variables. A p-value less or equal to 0.05 was considered statistically significant.

St. Mary’s Hospital, Lacor Institutional, Ethics, and Review Committee (Lacor IREC) approved the study. We obtained administrative clearance from the health facilities and informed consent from each participant. The research team ensured confidentiality of personal information during and after the investigation, and we only retained unique identifiers of participants on public records. In addition, only the principal investigator had access to the database during and after the project, which was archived at the Gulu University, Faculty of Medicine, Department of Surgery.

## Results

The prevalence of health insurance coverage among the study population was low, 57/723(7.9%), with most insured, 42 out of 57(73.7%) accepted the COVID-19 vaccine with a mean age of 33.81 years SD+8.863 at 95% CI:31.46-36.16. The uninsured but accepted the COVID-19 vaccine were 538/723(74.4%) and younger, with a mean age of 31.15 years SD+10.149 at 95% CI:30.38-31.92 and a median age of 29 years. The insured and uninsured ages range between 18-52 years and 18-75 years, respectively. The Likelihood ratio for COVID-19 vaccine acceptance with the insured participants was among age groups 9.813; df=4; p=0.044.

Widows, divorcees, and marriage separate participants, those from the remote districts of northern Uganda (Nwoya and Lamwo) and those with no formal education had no health insurance coverage. A multivariable logistic regression analysis showed that health insurance coverage among participants was not an independent predictor of COVID-19 vaccine acceptance AoR=1.304, 95%CI:0.657-2.732; p=0.421.

Table 1 shows participants with health insurance coverage and accepted the COVID-19 vaccine, with a prevalence rate of 42/57(73.7%), the mean age of 33.81 years SD+8.863 at 95% CI:31.46-36.16 and a median age of 35 years. Those without insurance but accepted the COVID-19 vaccine were 538/723(74.4%), and the mean age was 31.15 years SD+10.149 at 95% CI:30.38-31.92 and a median of 29 years. The insured and uninsured ages ranged between 18-52 years and 18-75 years, respectively.

**Table 1:** **Health Insurance Coverage and acceptance of the COVID-19 vaccine among participants in northern Uganda**.

| s/no | Variables | Yes (%) | No (%) |
| --- | --- | --- | --- |
| 1 | Health Insurance Coverage (HIC) | 42/723(5.8%) | 538/723(74.4%) |
| 2 | Mean age (years) | 33.81 | 31.15 |
| 3 | The standard error (SE) | 1.174 | 0.393 |
| 4 | 95% Confidence Interval (CI) | 31.46-36.16 | 30.38-31.92 |
| 5 | Median | 35.00 | 29.00 |
| 6 | Variance | 78.551 | 103.012 |
| 7 | Standard Deviation (SD) | 8.863 | 10.149 |
| 8 | Minimum | 18.00 | 18.00 |
| 9 | Maximum | 52.00 | 75.00 |
| 10 | Range | 34.00 | 57.00 |
| 11 | Interquartile range | 13.00 | 14.00 |
| 12 | Skewness | 0.167(SE=0.316) | 1.118(SE=0.095) |
| 13 | Kurtosis | -0.797 (SE=0.623) | 1.325(SE=0.189) |

Table 1 shows the participants with health insurance coverage that accepted the COVID-19 vaccine, with a prevalence of 42/723(5.8%), the mean age of 33.81 years SD+8.863 at 95% CI:31.46-36.16 and a median age of 35 years. Those without insurance cover but accepted the COVID-19 vaccine were 538/723(74.4%), and the mean age was 31.15 years SD+10.149 at 95% CI:30.38-31.92 and a median age of 29 years. The age ranges for the insured and uninsured were 18-52 years and 18-75 years, respectively.

Figure 1 is a box plot showing ages, COVID-19 vaccine acceptance, and health insurance cover among participants. Insured participants had an older average age (33.84 years) compared to those without 31.15 years.

**Figure 1:**
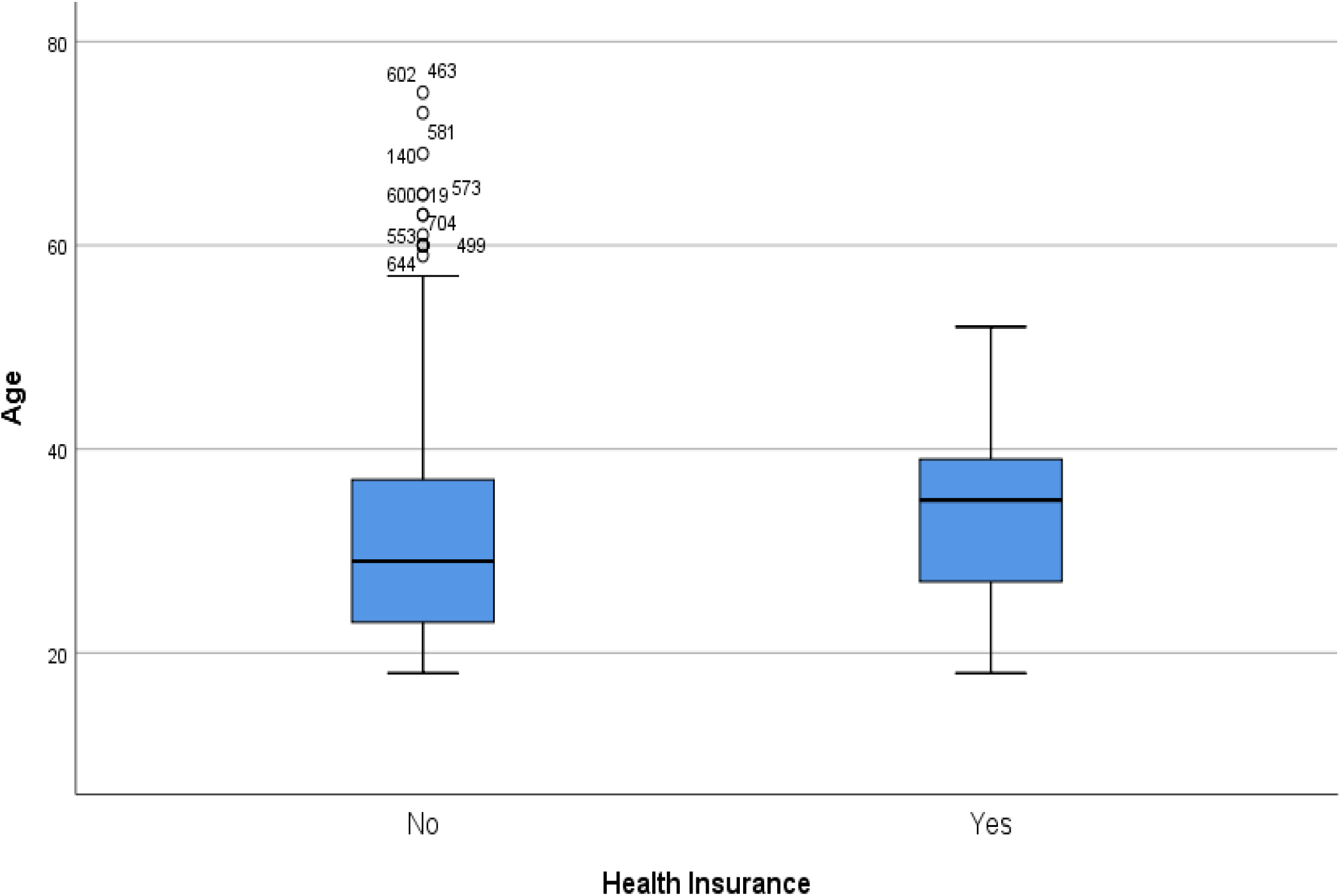
The ages, COVID-19 vaccine acceptance, and health insurance coverage among participants. Figure 1 is a box plot showing ages, COVID-19 vaccine acceptance, and health insurance cover among participants. Insured participants had an older average age (33.84 years) compared to those without 31.15 years.

Table 2 shows the association factors between health insurance coverage and other socio-demographic characteristics. Near significant association was observed with age groups χ^2^=8.122; df=4; p=0.087; with a Likelihood ratio of 9.813; df=4; p=0.044. The most vulnerable participants, such as widows, divorcees, and marriage separate participants, had no one with health insurance coverage. In addition, participants from remote districts of northern Uganda (Nwoya and Lamwo, and those without formal education had no health insurance coverage.

**Table 2:**
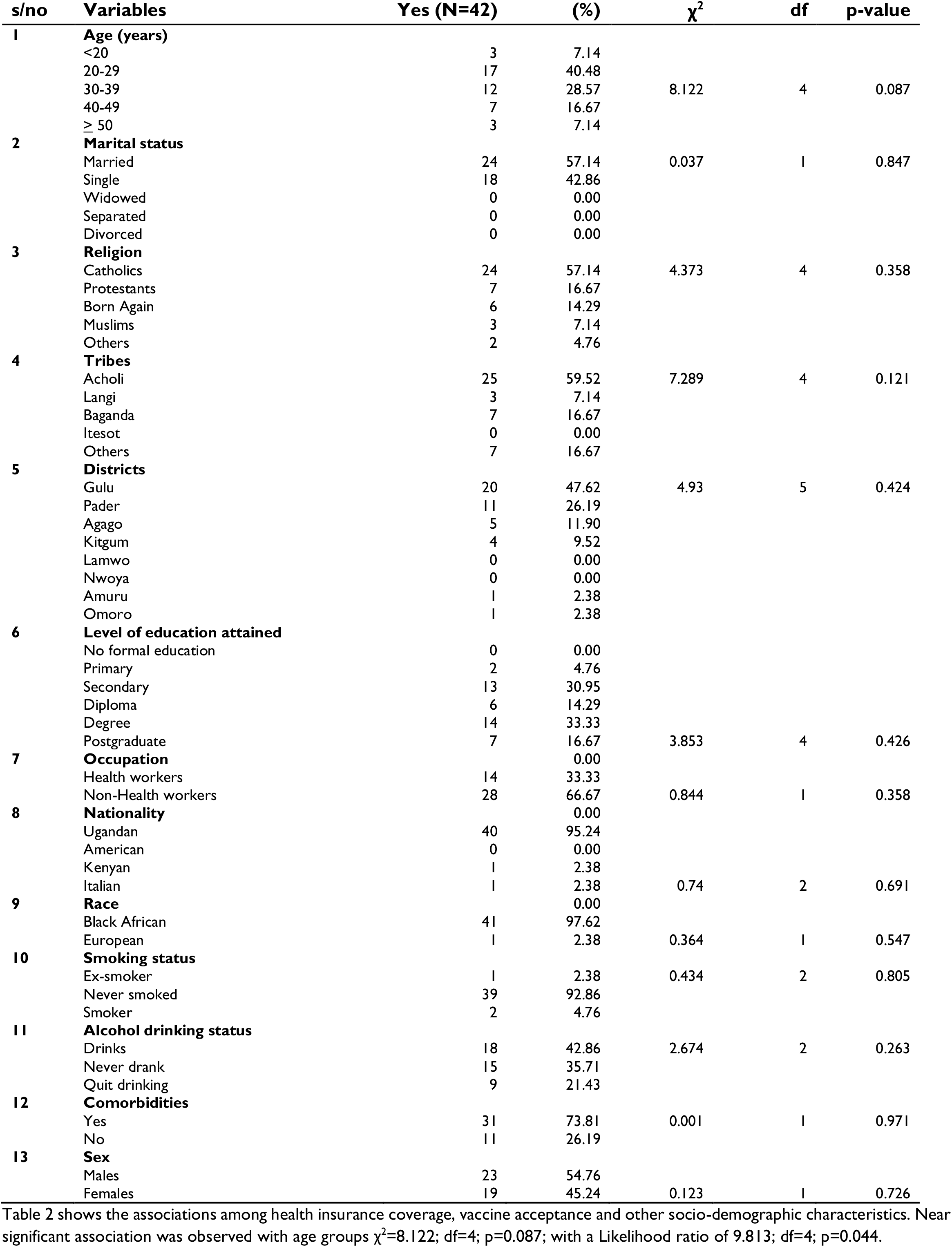
Health Insurance Coverage and COVID-19 vaccine acceptance among participants.

Table 3 shows the COVID-19 vaccine acceptance among the insured participants. COVID-19 vaccine acceptance among the insured participants was associated with no worries that they would take a COVID-19 vaccine by force χ^2^=8.036; df=1; p=0.005; did not get a fever after vaccination χ^2^=5.631; df=1; p=0.018; and mainstream media was not their most trusted sources of information on the COVID-19 χ^2^=4.619; df=1;p=0.032.

**Table 3:** Perceptions of COVID-19 among the insured participants in Northern Uganda.

| s/no | Variables | Yes (%) | No (%) | chi | df | p-values |
| --- | --- | --- | --- | --- | --- | --- |
| 1 | The mainstream media is my most trusted source of information | 15(35.7) | 27(64.3) | 4.619 | 1 | 0.032 |
| 2 | I got dizzy after receiving the COVID-19 vaccine | 2(4.8) | 40(95.2) | 0.740 | 1 | 0.390 |
| 3 | I got a blood clot after the COVID-19 vaccination | 7(16.7) | 35(83.3) | 0.916 | 1 | 0.339 |
| 4 | I have job-related worries with the COVID-19 pandemic | 9(21.4) | 33(78.6) | 3.487 | 1 | 0.062 |
| 5 | I am worried about the unavailability of the COVID-19 vaccine | 8(19.0) | 34(81.0) | 1.281 | 1 | 0.258 |
| 6 | I have food insecurity worries during this pandemic | 8(19.0) | 34(81.0) | 0.249 | 1 | 0.617 |
| 7 | The coronavirus is a plot or a conspiracy theory | 4(9.5) | 38(90.5) | 0.113 | 1 | 0.737 |
| 8 | I may be forced to take medicine for the virus | 0(0.0) | 42(100.0) | 2.850 | 1 | 0.091 |
| 9 | I may be forced to take the COVID-19 vaccine | 5(11.9) | 37(88.1) | 8.036 | 1 | 0.005 |
| 10 | I am not worried about any COVID-19 issues | 1(2.4) | 41(91.6) | 2.659 | 1 | 0.103 |
| 11 | I got a fever after the COVID-19 vaccination | 2(4.8) | 40(95.2) | 5.631 | 1 | 0.018 |

Table 3 shows the COVID-19 vaccine acceptance among insured participants was associated with no worries that they would be forced to take a COVID-19 vaccine $\chi^2=8.036$ ; $df=1$ ; $p=0.005$ ; did not get a fever after vaccination $\chi^2=5.631$ ; $df=1$ ; $p=0.018$ ; and mainstream media was not the most trusted source of information on the COVID-19 $\chi^2=4.619$ ; $df=1$ ; $p=0.032$ .

Health insurance coverage was not an independent predictor of COVID-19 vaccine acceptance at a multivariable logistic regression analysis AoR=1.501, 95%CI:0.807-2.791; p=0.199.

## Discussions

The most significant finding from this study was the low prevalence of health insurance coverage among the study population, 57/723(7.9%), with 42 of 57(73.7%) insured having accepted the COVID-19 vaccine with a mean age of 33.81 years SD±8.863 at 95% CI:31.46-36.16 (Table 1, Table 2). The insured but rejected the COVID-19 vaccine were 15 of 57 (26.3%). On the other hand, the uninsured but accepted the COVID-19 vaccine were 538/723(74.4%) and were younger, with a mean age of 31.15 years SD±10.149 at 95% CI:30.38-31.92 and a median age of 29 years (Figure 1 and Table 1). The insured participants and those without had ages ranging between 18-52 years and 18-75 years, respectively. The Likelihood ratio for COVID-19 vaccine acceptance among the insured was in the age group 9.813; df=4; p=0.044 (Table 1). The most vulnerable participants were widows, divorcees, and marriage separate participants who had no one with insurance coverage. In addition, participants from remote districts of northern Uganda and those without formal education had no insurance coverage too (Table 2). This finding has implications that during the COVID-19 pandemic, these vulnerable groups were adversely affected by the lack of access to health information and services because they had no health insurance coverage.

According to the WHO, Universal health coverage means that everyone has access to the health services they need, when and where they need them, without financial hardships [2]. It includes a full range of essential health services, from health promotion to prevention, treatment, rehabilitation, and palliative care [1,2]. In addition, Universal health coverage (UHC) ensures that people have access to the healthcare they need without suffering financial hardships [1,2,4]. UHC is also critical for achieving the World Bank Group’s (WBG) twin goals of ending extreme poverty and increasing equity and shared prosperity [1]. Ending extreme poverty and increasing equity are the driving forces behind the WBG’s health and nutrition investments [1].

Currently, at least half of the world’s population does not receive the health services they need [1,4,11,12]. About 100 million people are pushed into extreme poverty each year because of out-of-pocket spending on health [1,4]. To make health a reality for all, individuals, and communities with access to high-quality health services for taking care of their health and the health of their families are necessary. Skilled health workers provide quality, people-centered care, and policymakers are committed to investing in universal health coverage [1,2,12]. Universal health coverage must be strong, people-centered primary health care with sound health systems rooted in the communities they serve [1,11,12]. They focus not only on preventing and treating diseases and illnesses but on helping to improve the general well-being and quality of life [1,2].

Our study found that the most vulnerable participants, such as widows, divorcees, and married separate, had no health insurance coverage (Table 2). In addition, participants from remote districts of northern Uganda and those without formal education were not insured (Table 2). The lack of health insurance coverage among most of the study participants has implications. During the COVID-19 pandemic, the population was adversely affected by a lack of access to health information and services, usually available to those with health insurance coverage.

Interestingly, discussions on the national health insurance scheme in Uganda started in the late 1980s and evolved slowly over the years [3]. In 2017, discussions on the plan accelerated when Advance Family Planning’s local partner, PPD ARO, stakeholders, Population Action International (PAI), and The William and Flora Hewlett Foundation worked to ensure that the draft scheme had family planning programs [3]. The coalition steered advocacy efforts to provide family planning commodities and services that the stakeholders formed throughout the scheme’s development processes [3]. Between 2016 and 2020, Parliamentary champions asked PPD ARO to organize a series of meetings with members of Parliament, the Ministry of Health, the Ministry of Finance, and the Uganda Reproductive, Maternal, Newborn, Child, and Adolescent Health (RMNCAH+N) civil society platform to continue this momentum [3]. For three consecutive years, Uganda’s representatives in the Network of African Parliamentary Committees of health, a regional entity, committed to passing the national health insurance scheme but struggled to get the Ugandan Ministry of Health to draft and introduce the bill to Parliament [3]. However, in June 2019, Parliamentary champions sensed a stalling of the draft bill in the Ministry of Health [3]. They asked PPD ARO for technical support to inform a private member’s account for Hon. Dr. Michael Bukenya, the chairperson of the parliamentary health committee [3]. The introduction of the private member’s bill put pressure on the government to revive, update, and present their bill to Parliament in August 2019. Eventually, the Ugandan Ministry of Health’s revised bill incorporated elements from the private member’s bill in its final form [3].

Therefore, in February 2020, PPD ARO published an issue brief on the draft scheme and organized a series of media engagement activities to share details and benefits of the plan with the public, including its coverage of family planning [3]. The advocacy kept the issue central to parliamentarians all over the country as the COVID-19 pandemic unfolded and challenged national health financing [3]. The media were instrumental in keeping the pressure on Parliament to pass the bill, which finally happened on March 31, 2021 [3]. However, this bill has not been assented to and has effectively prevented the population from accessing services as required by the UHC declarations and SDGs [3].

Interestingly, every country worldwide has committed to achieving universal health coverage (UHC) by 2030 as part of the United Nations Sustainable Development Goals (SDGs) [4,11,12]. But, some countries are progressing faster than others in delivering equitable access to health services, affordable medicines, and vaccines [11].

Among those leading the pack is Vietnam. Today, 87.7% of Vietnam’s population, or 83.6 million people, are covered by health insurance [11]. According to the latest Global Monitoring Report on UHC, published jointly by the World Health Organization and the World Bank, 97% of Vietnamese children now receive standard immunizations, compared to 95% of children in the United States [11]. Since 1990, the country’s maternal mortality rate has fallen by 75% [11]. Vietnam has reached such impressive milestones ahead of schedule, despite having an average per capita income of just $2,342 as of 2017 [11]. The key to its success is not the scale of investment in healthcare, which amounts to a modest $142 per person annually (including both public funding and out-of-pocket expenses), but rather how the government uses its resources, including the country’s intellectual capital [11].

Vietnam’s strategic approach was in its Ministry of Health’s directing healthcare activities scheme, which required health facilities at the central and provincial levels of government administration to help build up the capacity of district and community facilities [11]. A key objective of this scheme was to shift more of the burden of delivering medical services from higher-level hospitals onto lower-level primary healthcare facilities [11]. Given a long history of deep disparities in health outcomes between urban and rural areas, Vietnamese still often tried to bypass their local healthcare centers in favor of major hospitals in urban centers [11]. This created inefficiencies in the health system and increased out-of-pocket costs for patients and their families without guaranteeing the best care [11].

Thus, going beyond ensuring that community health facilities can offer affordable, quality care, there is a need to change public perceptions [11]. Families need to trust that they can get a dependable diagnosis of malaria, chronic obstructive pulmonary disease, or diabetes locally, as well as the necessary medications and other treatments. Therefore, health facilities must strengthen their relationships with local communities by routinely providing a level of service that satisfies patients [11]. Such connections will help to advance another health-improving, cost-saving imperatives where local health workers should be able to educate their communities to maintain health and avoid illnesses. Success will require good working conditions and access to the ongoing training and management support that are critical to job satisfaction [11].

Vietnam’s government recognized that to implement its healthcare strategy effectively, it needed help from other partners [11]. It then established a Working Group for primary healthcare transformation led by the Vietnamese Ministry of Health and included diverse actors from the public, nonprofit, and private sectors [11]. The group’s founding partners were the World Economic Forum, Harvard Medical School, and Novartis [11]. The working group aimed to strengthen existing primary care demonstration projects in 30 Vietnamese provinces and applied the lessons learned to develop holistic solutions that could be replicated and scaled up in Vietnam [11]. It also prioritized rigorous measurements and evaluation of outputs, from the quality of community-level health services to the cost-effectiveness of primary health care [11]. The government invited each partner to contribute capabilities, resources, and knowledge to this endeavor. For example, Harvard Medical School brought world-class expertise in the organizational management of primary healthcare teams [11]. National partners brought, among others, a deep understanding of the local context, which was essential for developing and implementing sustainable solutions [11]. For its part, Novartis offered insight on how to deploy digital technology at a large scale, engage rural communities in health education, and expand education programs for healthcare practitioners in rural communities [11]. Notably, Novartis made similar contributions through another successful public-private partnership in Vietnam, Cùng Sông Khòe [11].

In partnership with Vietnam’s government, Cùng Sông Khòe (CSK) has been delivering services to underserved rural communities in Vietnam since 2012 [11]. That initiative expanded healthcare for common medical conditions like diabetes, hypertension, and respiratory diseases, patient health education, and continuing medical education for health professionals [11]. Since 2012, CSK has reached out to more than 570,000 people, mainly adults, across the sixteen provinces of Vietnam [11].

It would therefore be naïve to think that all going on in Vietnam are beds of roses because Vietnam is facing significant challenges ahead [11]. It is grappling with behavioral and environmental factors underlying poor health and diseases, especially high rates of smoking among males, high rates of alcohol consumption, air pollution, and the rapidly growing number of aging populations [11]. Governments must conduct critical healthcare reforms to improve healthcare outcomes; for example, the government should incentivize doctors to be more selective in referring patients to higher-level hospitals and sending more patients to local primary-health-care centers [11]. Nonetheless, Vietnam’s progress toward UHC has been remarkable, partly due to the government’s embrace of strategic public-private partnerships (PPP) [11]. For countries that have struggled to move forward, this model and approaches from other high performers in the race for UHC, such as Indonesia, Rwanda, and Thailand, may be worth embracing [11].

### The global movement toward Universal Healthcare Coverage (UHC)

Health is an essential part of the Sustainable Development Goals (SDGs). For example, the SDG 3.8 target aims to achieve universal health coverage, including financial risk protection, access to quality essential healthcare services, and safe, adequate, quality, and affordable essential medicines and vaccines for all [1,2,4,11,12]. In addition, SDG 1, which calls to “end poverty in all its forms everywhere, could be in peril without UHC, as almost 90 million people worldwide are impoverished by health expenses every year [4,11,12]. Access to affordable, quality primary healthcare is the cornerstone of UHC, but many people worldwide still struggle to fulfill their immediate healthcare needs [11,12,1,13,14]. Often overlooked, mental health is also an essential element of UHC, as it is critical to people’s ability to lead productive lives [11].

In recent years, the UHC movement has gained global momentum, with the first-ever UN high-level meeting on UHC held in September 2019 in New York, USA [4,11]. Member states unanimously adopted a Political Declaration, affirming their high-level political commitment to UHC and outlining several necessary actions [11]. Twelve co-signatories, including the WBG, also launched the Global Action Plan (GAP) for healthy lives and well-being to support countries in jointly delivering on the SDG3 targets [11]. In January 2020, the second UHC forum was held in Bangkok to enhance political momentum on UHC in international outlets [11].

### Providing affordable and quality primary healthcare

Providing affordable, quality health services to the community, women, children, adolescents, and people affected by mental health issues represents a long-term investment in human capital [11,14,15]. Primary health services are a fundamental element of UHC, yet research warns that, if current trends continue, up to 5 billion people will still be unable to access health care in 2030 [14]. Maternal and child mortality remains high in several parts of the world. More than a fourth of girls and women in sub-Saharan Africa cannot access family planning services, encouraging unplanned pregnancies and maternal, infant, and child mortality and morbidity [14,15].

In 2015, the WBG and partners set up the Global Financing Facility (GFF), a multi-stakeholder initiative that focuses on helping countries improved maternal, child, and adolescent health services [15,16]. Many countries experiencing rapid population growth have young populations that could drive economic growth and reduce poverty [15,16,17]. But to unleash the benefits of the demographic dividend, governments must invest in the health and well-being of their people to build human capital and boost inclusive growth [15,16,17,18]. Improving reproductive, maternal, newborn, child, and adolescent health (RMNCAH) and addressing mental health disorders are crucial for achieving UHC because significant challenges exist.

### Maternal mortality

Most of the world’s maternal deaths occur in developing regions; in the least developed countries, the lifetime risk of maternal death for women is, on average, one in fifty-six compared to one in 7,800 in high-income countries like Australia or New Zealand [14,15,16]. In sub-Saharan Africa, which accounts for two in three maternal deaths (66%), the risk is one in 37 [15,16]. A further 20% of maternal deaths occur in South Asia, and most of these fatalities are preventable if pregnant women have timely access to the necessary healthcare [16,17].

### Child mortality

Reports show that mortality rates among children under five have more than halved from 12.5 million to 5.2 million between 1990 and 2018, according to a joint 2020 report published by the WBG, WHO, and UNICEF [17]. Yet a child’s chance of survival depends on where they are born [17]. Worldwide, 15,000 children under five still die every day [17]. In sub-Saharan Africa, one child in 13 dies before their fifth birthday compared to one in 199 in high-income countries [17,18].

The WBG, WHO, and UNICEF also collaborated on another 2020 publication that highlighted stillbirths, an issue that remains largely overlooked [18]. Every year, 2 million babies are stillborn worldwide, and progress in reducing these numbers has not kept up with the decline in under-five mortality [18]. In 2000, the ratio of stillbirths to under-five deaths was 0.30, but by 2019, it had risen to 0.38 worldwide. In sub-Saharan Africa, stillbirths increased from 0.77 million in 2000 to 0.82 million in 2019 [18].

### High fertility rates

Globally, women are giving birth to fewer children today than three decades ago [19]. However, there are still a handful of countries with persistently high fertility rates, such as Niger (7.0), Mali (6.0), and the Democratic Republic of Congo (6.0) [19]. In countries with lower fertility, such as Ethiopia, fertility varies within different regions [19]. It ranges from 1.7 in Addis Ababa, the capital city, to 6.4 in Somali, a regional state [19]. Countries with persistently high fertility often face high maternal, infant, and child mortality burdens [19].

### Adolescent fertility

More teenage girls are giving birth in countries with high fertility rates [19,20]. In the sub-Saharan Africa, the adolescent fertility rate is 102 births per 1,000 girls [19,20,21,22]. Underage mothers are more likely to experience complications due to pregnancy, such as obstructed labor and eclampsia, increasing their risks of death [20,21,22,23,24]. In addition, children born to adolescents are also more likely to have a low birth weight, ill-health, stunting, and other poor nutritional outcomes [19,20-25].

### Mental, neurological, and substance use disorders (MNS)

These are common, highly disabling disorders that are associated with significant premature mortality, and they impose a human, social and economic toll [26-27]. Every 40 seconds, a person commits suicide worldwide [26-27]. Therefore, to fully realize the goal of universal health coverage and improve human capital outcomes worldwide, mental health programs must be integrated with service delivery at the community level and covered under financial protection arrangements [26,27]. Estimates suggest that nearly 1 billion people live with a mental disorder worldwide [26,27]. In low-income countries, more than 75% of people with the disease do not receive treatment, and approximately half of all mental health disorders emerge by age 14 and some other illnesses [26,27]. More than one in five people (22.1%) suffer from mental ill-health in countries affected by fragility, conflict, and violence [26,27]. Women and children who have experienced violence, soldiers returning from war, migrants and refugees displaced by conflict, the poor, and other vulnerable groups in society are disproportionately affected [26,27]. The COVID-19 pandemic has caused a global increase in mental health disorders due to various factors, including anxiety, lockdowns, and job losses while disrupting or halting critical mental health services in many African countries [28,29]. Since MSN have an early onset, often in childhood or early adolescence, and are highly prevalent in the working-age population, they contribute to economic output losses estimated between $2.5-8.5 trillion globally. The World Bank and World Health Organization project that mental health issues will double by 2030 [26,27].

### Mobilizing resources for UHC

In June 2019, the President of Japan hosted the first-ever G20 Finance and Health Ministers joint session on resource mobilization for UHC [30]. The discussion aimed to galvanize G20 countries towards the common goal of financing UHC in developing countries [30]. A World Bank report showed that people in developing countries spend half a trillion dollars annually, over $80 per person, out of their own pockets to access health services [30]. Such expenses hit the poor the hardest and threatened decades-long progress in health [30].

World Bank/World Health Organization (WHO) research from 2019 shows that countries must increase spending on primary health care by at least 1% of their gross domestic product (GDP) if the world is to close glaring coverage gaps and meet the health targets agreed under the SDGs [30]. A lack of universal access to quality, affordable health services endanger countries’ long-term economic prospects and make them more vulnerable to pandemic risks [30]. Developing countries, faced with a growing number of aging populations and burdens of non-communicable diseases, need urgent action. They find themselves increasingly challenged to close the gap between the demand for health spending and available public resources, which prolongs patients’ families’ reliance on out-of-pocket expenditures [30].

### Universal Health Coverage for inclusive and sustainable development

In 2011, Japan celebrated the 50^th^ anniversary of its achievement of universal health coverage (UHC) [12-15]. On this occasion, the government of Japan and the World Bank Group decided to undertake a multi-country study to share varied experiences from countries at different stages of adopting and implementing strategies for UHC, including Japan itself [12-16]. The initiative resulted in an in-depth report on Japan’s experience entitled “Universal Health Coverage for Inclusive and Sustainable Development: Lessons from Japan” [12,13]. The goals of UHC are to ensure that all people can access quality health services. To safeguard all people from public health risks, and protect people from impoverishment due to illness, whether from out-of-pocket payments for healthcare or loss of income when a household member falls sick [12,31-36].

Countries as diverse as Brazil, France, Japan, Thailand, and Turkey have shown how UHC can serve as a vital mechanism for improving the health and welfare of their citizens, as well as lay the foundation for economic growth grounded in the principles of equity and sustainability [12,31-37]. Ensuring universal access to affordable, quality health services will end extreme poverty by 2030 and boost shared prosperity in low- and middle-income countries, where most of the world’s poor reside [12,38].

While governments can, and should, play a leading role in the global UHC movement, to make the dream of UHC for all a reality, governments, nonprofit organizations, and businesses must work together to create and invest in robust health systems [12,13,31-40]. Global Citizen and Johnson & Johnson support the UN Sustainable Development Goal of ensuring people’s healthy lives and well-being are taken care of no matter who they are, where they live, or their income [11,12].

In Uganda, the Parliament has passed the national health insurance scheme, but more advocacy is needed to ensure that the President signs it promptly [3]. However, private employers have publicly opposed the plan, fearing that paying employees’ contributions would raise business costs [3]. The scheme advocates need to continue to engage the President to highlight the benefits of the national health insurance scheme [3]. They should plan to maintain pressure to sign the bill by continuing media coverage and strategic messaging [3]. Furthermore, civil society champions should remain engaged when the bill becomes law and support the scheme’s regulations process and implementation [3].

In summary, the study found that Ugandans’ health insurance coverage is low, with the most vulnerable population such as divorcees, widows, marriage separate participants, those without formal education, and from remote districts of northern Uganda were the most affected. The problem Uganda is experiencing is not the unaffordability of the COVID-19 vaccine and its related costs because vaccines are offered free and at the nearest health facility by the Government of Uganda. It is instead the access to information and cost of treatment when one is affected by COVID-19 that are not readily available and affordable to most Ugandans because of the lack of national health Insurance coverage. Also, when vaccinated, the fear of vaccine side effects, complications, and where to find remedies have created vaccine hesitancy/ inquisitiveness among the population. Whether true or false, these perceptions are the reality in Ugandan communities that the Government of Uganda must handle. Therefore, if Uganda’s national health insurance coverage bill became law, access to health information and treatment for COVID-19 would have been available and open to the general population, especially those without health insurance coverage.

So, the COVID-19 pandemic exposed Uganda’s health service delivery vulnerability, which should provide a blueprint for future epidemic preparedness. The need for a law on Uganda’s national health insurance coverage has become necessary for this country if we are to achieve Universal Health Coverage (UHC) and Sustainable development goals (SDGs) soon.

### Strengths and Limitations

The number of participants with health insurance coverage was small, which limited us from performing some of the analysis as some cells did not have enough numbers. A more extensive sample survey including many regions of Uganda would provide the power and accuracy of findings. However, this data is vital as it is one of the well-documented and completed data for over 723 participants from the Acholi sub-region regarding COVID-19 vaccine acceptance in the recent period. Findings from this study show a high acceptance rate of the COVID-19 vaccine, especially among the insured, despite results from other parts of Uganda.

### Generalizability of results

Findings from this study should be interpreted cautiously in regions with low-resource settings in Uganda.

## Conclusions

As the world grapples with the control of COVID-19, vaccine acceptance and health insurance coverage have become critical issues to be handled by each country. The health insurance coverage among participants from northern Uganda was low at 57/723(7.9%). Most participants with health insurance coverage accepted the COVID-19 vaccines compared to those who did not. The lack of health insurance coverage among most study participants is problematic as the world looks toward attaining UHC and SDGs. We propose that Uganda’s national social health insurance scheme, which is not law, is urgently reviewed and signed to allow Uganda’s population access to the needed health services.

## Data Availability

All datasets supporting this article's conclusion are within the paper and are accessible by a reasonable request to the corresponding author.

## Declarations

Ethics approval and consent to participate: The St. Mary’s Lacor Hospital Institutional and Ethics Committee (LHIREC) approved this study. In addition, the study followed the relevant institutional guidelines and regulations.

## Authors’ contributions

DLK, ENI, JA and FWDO participated in designing the study, JNO, LO, FWDO, and DLK were responsible for supervising data collection, LO and DLK were responsible for data analysis and interpretation, CO, DA, JNO, ENI, FWDO, LO, JA, DLK for writing and revising the manuscript. All Authors approved the manuscript.

## Authors’ Information

Dr. Eric Nzirakaindi Ikoona (ENI) is a Technical Director at ICAP at the University of Columbia, Sierra Leone; Dr. Freddy Wathum Drinkwater Oyat (FWDO) is a senior physician, a public health specialist, and a member of Uganda Medical Association, UMA-Acholi branch, Gulu City, Uganda; Mr. Lawrence Oballim is a member of Gulu University, Faculty of Science, Department of Computer Science, Gulu City, Uganda; Dr. Judith Aloyo (JA) is a Technical Director at the Rhites-N, Acholi, Gulu City, Uganda; Prof. David Lagoro Kitara (DLK) is a Takemi fellow of Harvard University and a Professor at Gulu University, Faculty of Medicine, Department of Surgery, Gulu City, Uganda

## Acknowledgment

We acknowledge with many thanks for assistance from the administration of health facilities in the region for the information obtained. Financial support from UMA Acholi branch members, which enabled the team to conduct this study successfully, is most appreciated.

